# PSEN1 loss-of-function variants do not cause early-onset Alzheimer’s disease

**DOI:** 10.64898/2026.11.13.26367865

**Authors:** Amelia Farinas, Andrés Peña-Tauber, Timothy J. Hohman, Alex G. Contreras, Tanner D. Jensen, Yann Le Guen, Michael D. Greicius

## Abstract

*PSEN1* variants are the most common cause of familial early-onset Alzheimer’s disease (EOAD), but it is debated whether they act through a loss- or gain-of-function. Following a recent report of a duo with EOAD carrying a predicted loss-of-function (LoF) stop-gain variant (p.K109*) in *PSEN1*, we sought to determine whether *PSEN1* LoF variants, as a class, cause EOAD. We identified 45 carriers of *PSEN1* LoF variants predicted to trigger nonsense-mediated decay in the Alzheimer’s Disease Sequencing Project, UK Biobank, and BioVU databases. None of these carriers had a recorded diagnosis of early-onset dementia, and 17 were older than 65 years at last follow-up with no dementia or AD reported. Re-analysis of the reported AD-associated variant p.K109* revealed a cryptic splice site predicted to produce an in-frame deletion of 5 amino acids (p.K109_L113del). These results suggest that *PSEN1* variants must produce an altered PSEN1 protein product to cause EOAD, while true null LoF *PSEN1* alleles do not cause EOAD.

## Main Text

The presenilin proteins are central in Alzheimer’s disease (AD). Together, *PSEN1* and *PSEN2* contain most of the identified mutations in familial AD (FAD), and their protein products serve as the catalytic subunits of the γ-secretase complex responsible for amyloid precursor protein (APP) cleavage and amyloid-β (Aβ) production^1–3^. Most pathogenic *PSEN1* variants associated with FAD are missense mutations, with typical ages of onset ranging from 30-50 years and near complete penetrance^4^. However, it remains debated whether abnormal presenilin activity in AD is best understood as a gain of toxic function (GoF) or a loss of normal function (LoF). This ambiguity has hindered our mechanistic understanding of AD and the development of effective therapeutics.

Because cleavage of APP by γ-secretase produces Aβ, proponents of the amyloid hypothesis of AD initially argued that *PSEN1* mutations were gain-of-function, increasing the production of aggregation-prone Aβ42^5,6^. However, studies of additional FAD *PSEN1* mutations demonstrated that many actually impaired γ-secretase-dependent APP processing, decreasing production of both Aβ42 and Aβ40^7^. The ratio of Aβ42/Aβ40, or long to short Aβ products more generally, can be a strong predictor of mutation age-at-onset in FAD^8,9^; this supports the idea that *PSEN1* mutations may drive disease by altering the enzymatic properties of γ-secretase, for example by shifting production towards longer Aβ peptides. However, it remains unclear whether these changes in Aβ processing are best understood as a gain or loss of *PSEN1* function.

More than 300 *PSEN1* mutations, present at more than 20% of its amino acid residues, have been linked to FAD. This diffuse distribution is evocative of a LoF mechanism^10^. PSEN1 proteins with FAD mutations show reduced ability to rescue the effects of *PSEN1*-homolog knockouts in *C. elegans* and *Drosophila*, suggesting impairment of endogenous *PSEN1* function^1^. Mice with conditional *PSEN1* deletion show progressive memory loss, synaptic and neuronal loss, astrogliosis, and tau hyperphosphorylation without amyloid accumulation^1^, linking *PSEN1* loss of function to neurodegeneration independent from effects on amyloid processing. Loss of amyloid-independent *PSEN1* functions like Notch cleavage, calcium signaling, autophagy, or lysosomal homeostasis have thus been proposed to play a role in FAD^11^. However, high-quality reports of variants resulting in gene-level *PSEN1* LoF in AD are limited, despite reports linking these mutations to other phenotypes related to loss of γ-secretase function^12–15^.

A recent report of a father-son duo with early-onset AD (EOAD) harboring a stop-gain variant (K109*) in *PSEN1* renewed interest in this critical question and provided some support for the possibility that *PSEN1* mutations causing EOAD reflect a loss-of-function mechanism. Both father and son were heterozygous carriers of the K109* variant; the son developed dementia with amyloid-positive PET in his 30s, and the father had dementia in his 40s, with biopsy-confirmed pathology consistent with Alzheimer’s disease. After functional characterization of the K109* variant in cell and mouse models, the extent to which this variant caused the duo’s disease, and its potential mechanism, remained unclear^16^.

To understand whether gene-level *PSEN1* LoF variants are causal for AD, we systematically searched for carriers of LoF variants in large clinical and population genetic databases: the Alzheimer’s Disease Sequencing Project (ADSP), which provides high-quality phenotyping for neurodegeneration including clinical and biomarker assessments, and population biobanks including UK Biobank (UKB), the All of Us Research Program (AoU), and Vanderbilt’s BioVU database, which together provide electronic health record data for over 1.1 million participants.

We identified 58 individuals in the ADSP, UKB, and BioVU databases with 42 unique variants annotated as high confidence loss-of-function variants by LOFTEE^17^ (Supplementary Tables 1 and 2). This included 6 splice variants affecting a splice donor or acceptor, and 36 frameshift and stop-gain variants. Two of the frameshift or stop-gain variants led to a premature stop in the last exon of *PSEN1*, likely resulting in a truncated protein that escapes nonsense-mediated decay^18^. Additionally, splice variants may lead to production of an altered protein that otherwise retains the wild-type reading frame^19^. To focus on true genetic LoF mutations, in which no or minimal protein is produced by the mutant allele, we considered only the 34 frameshift or stop-gain mutations that were predicted to lead to nonsense-mediated decay as high-confidence LoF mutations.

Of the 45 individuals carrying high-confidence *PSEN1* LoF mutations, none had early-onset dementia. 42 individuals remained dementia-free through their last assessment, including 17 carriers who were observed past age 65 (Figure 1). Although 3 individuals had diagnoses of dementia past the age of 65, their onset ages are more similar to individuals with sporadic, late-onset dementia than the early onset observed in carriers of familial *PSEN1* variants. We also identified fewer than 20 carriers of *PSEN1* LoF mutations in the AoU database; while specific findings could not be disclosed under AoU data dissemination policies, the observed phenotypes were consistent with our conclusions. With the notable numbers of healthy, dementia-free individuals observed at older ages carrying high-confidence *PSEN1* LoF mutations, these mutations as a class are unlikely to cause FAD.

**Figure 1:**
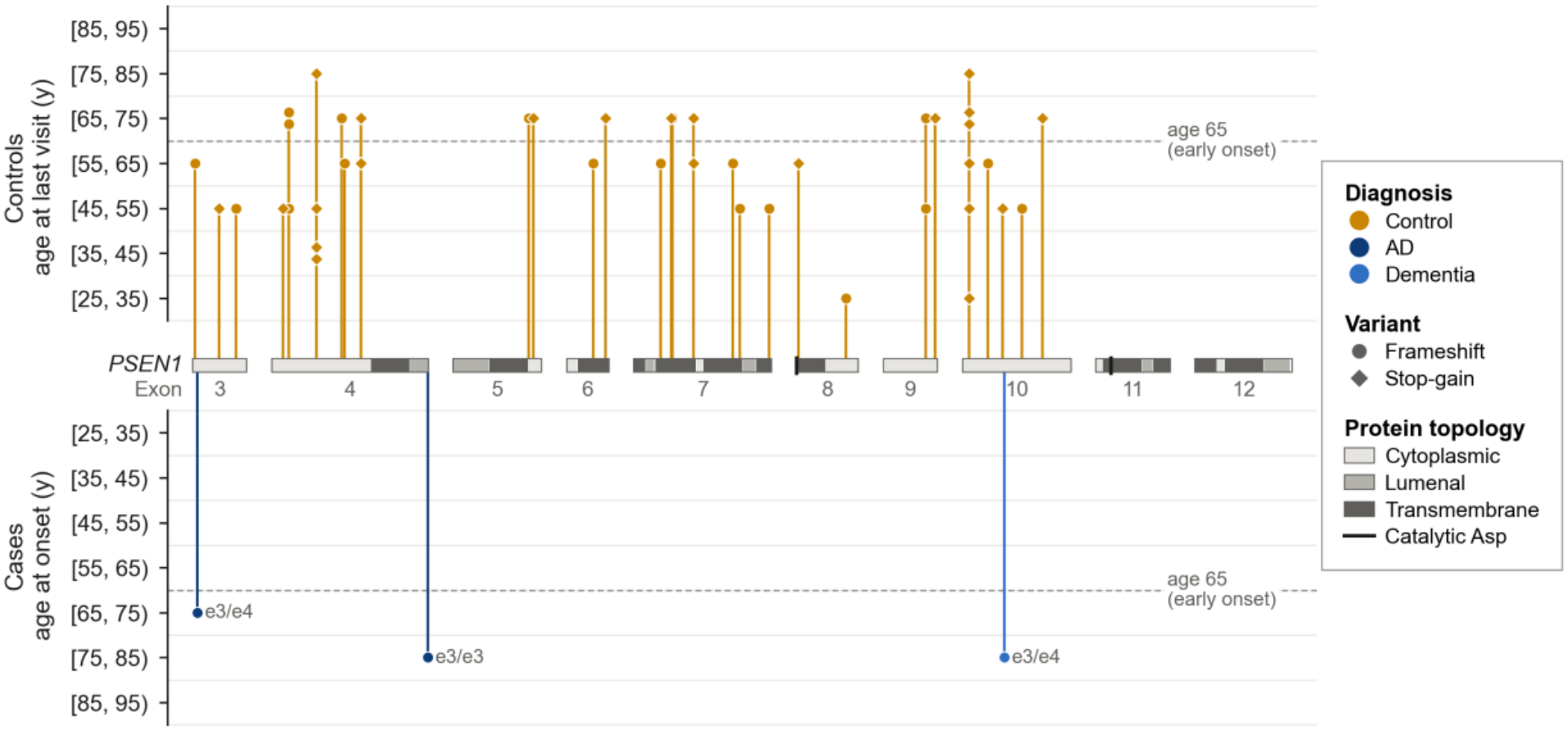
*PSEN1* high confidence LoF mutation carriers in ADSP, BioVU, and UKB. Each marker represents one carrier (n = 45) of a frameshift or stop-gain *PSEN1* variant predicted to lead to NMD, drawn at the variant’s position along the *PSEN1* coding sequence. Controls (amber) are plotted above the gene at their age at last visit, and cases below the gene at their age at onset of AD (navy) or dementia (blue). Cases are labeled with their APOE genotype. Ages are grouped into 10-year bins to protect patient privacy.

We also examined the phenotypes of the 13 individuals carrying mutations that may lead to the production of alternatively spliced or truncated PSEN1 protein (Figure 2, Supplementary Table 3). Of these individuals, 6 had early-onset mild cognitive impairment (MCI) or EOAD, with disease onset before age 65. All early-onset cases were splice variant carriers at the exon 9 acceptor site (14:73206385:G:A and 14:73206385:G:T). Three carriers of the G>A variant were family members, as were two carriers of the G>T variant; the third G>T carrier was unrelated. These variants have been previously shown to cause familial AD. They result in in-frame skipping of exon 9 and produce a shortened, 438 amino acid protein product, with modified functions including amyloid processing and proteolytic cleavage^20^. Six other carriers of splice or truncation mutations had no diagnoses of MCI, AD, or dementia, while one individual, carrying a splice donor variant (14:73198130:G:C) predicted to result in in-frame skipping of exon 8, had MCI in her 70s. Thus, specific mutations that produce altered PSEN1 protein, including in-frame splice variants, may cause FAD, but these mutations are distinct from gene-level loss of function mutations.

**Figure 2:**
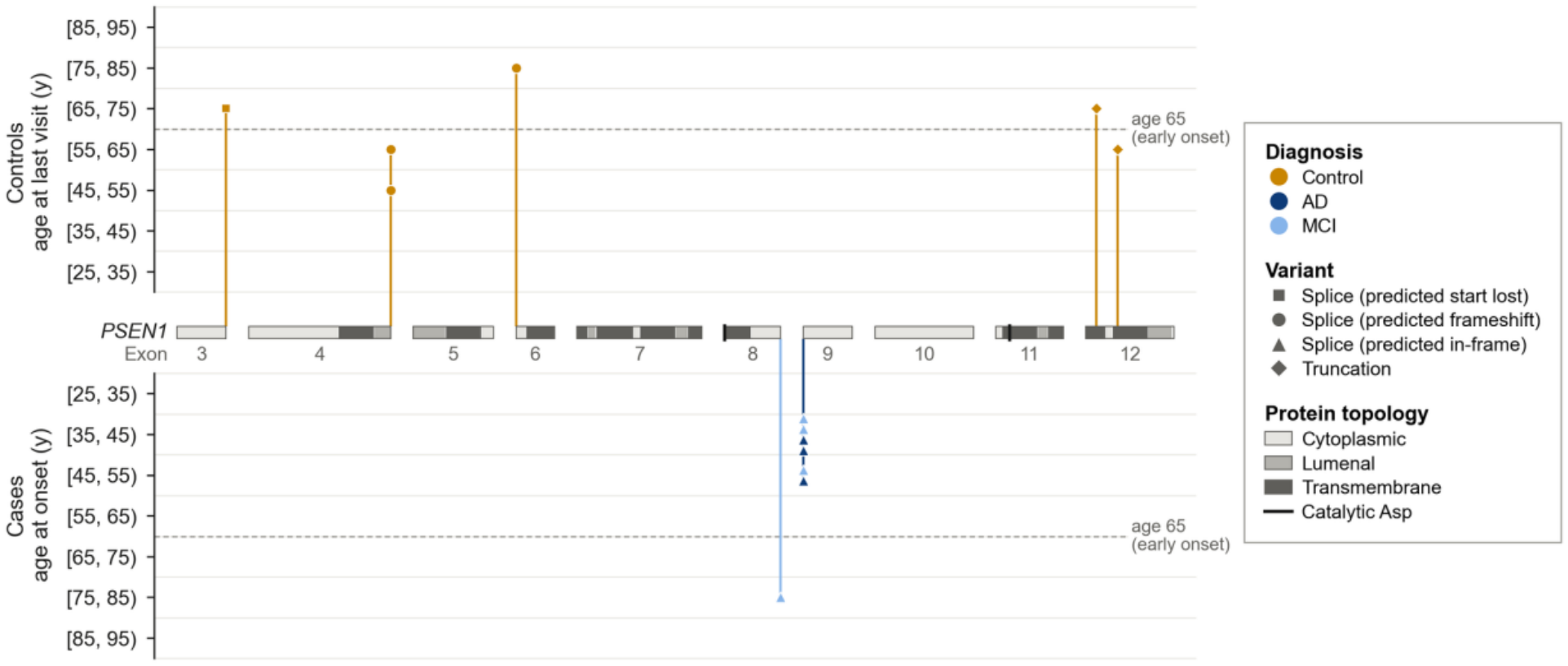
*PSEN1* splice and truncation mutation carriers in ADSP, BioVU, and UKB. Each marker represents one carrier (n = 13) of a *PSEN1* splice or truncation variant predicted as LoF by LOFTEE, drawn at the variant’s position along the *PSEN1* coding sequence. Controls (amber) are plotted above the gene at their age at last visit, and cases below the gene at their age at onset of AD (navy) or MCI (light blue). Ages are grouped into 10-year bins to protect patient privacy.

Given our finding that high confidence *PSEN1* LoF mutations are unlikely to cause EOAD, we revisited the reported case of the father-son duo with EOAD harboring the exon 4 stop-gain mutation (K109*). We searched the son’s genome for additional variants that could explain the phenotype but did not identify any additional rare variants likely to cause EOAD. As this duo’s clinical features were clearly reminiscent of *PSEN1* FAD, including onset in the 30s to 40s and substantial amyloid deposition in the basal ganglia^21^ observed in the son’s amyloid PET scan, we re-evaluated the K109* mutation (c.325A>T, Figure 3A) to ensure it was a true stop-gain loss of function mutation. Surprisingly, AlphaGenome predicted that the A>T mutation creates a cryptic splice site donor at c.323, 15 nucleotides before the canonical exon 4 splice donor at c.338 (Figure 3B). Three sequence-based splicing predictors all supported increased use of this cryptic donor and decreased use of the canonical exon 4 donor for the alternate allele (Figure 3C).

**Figure 3:**
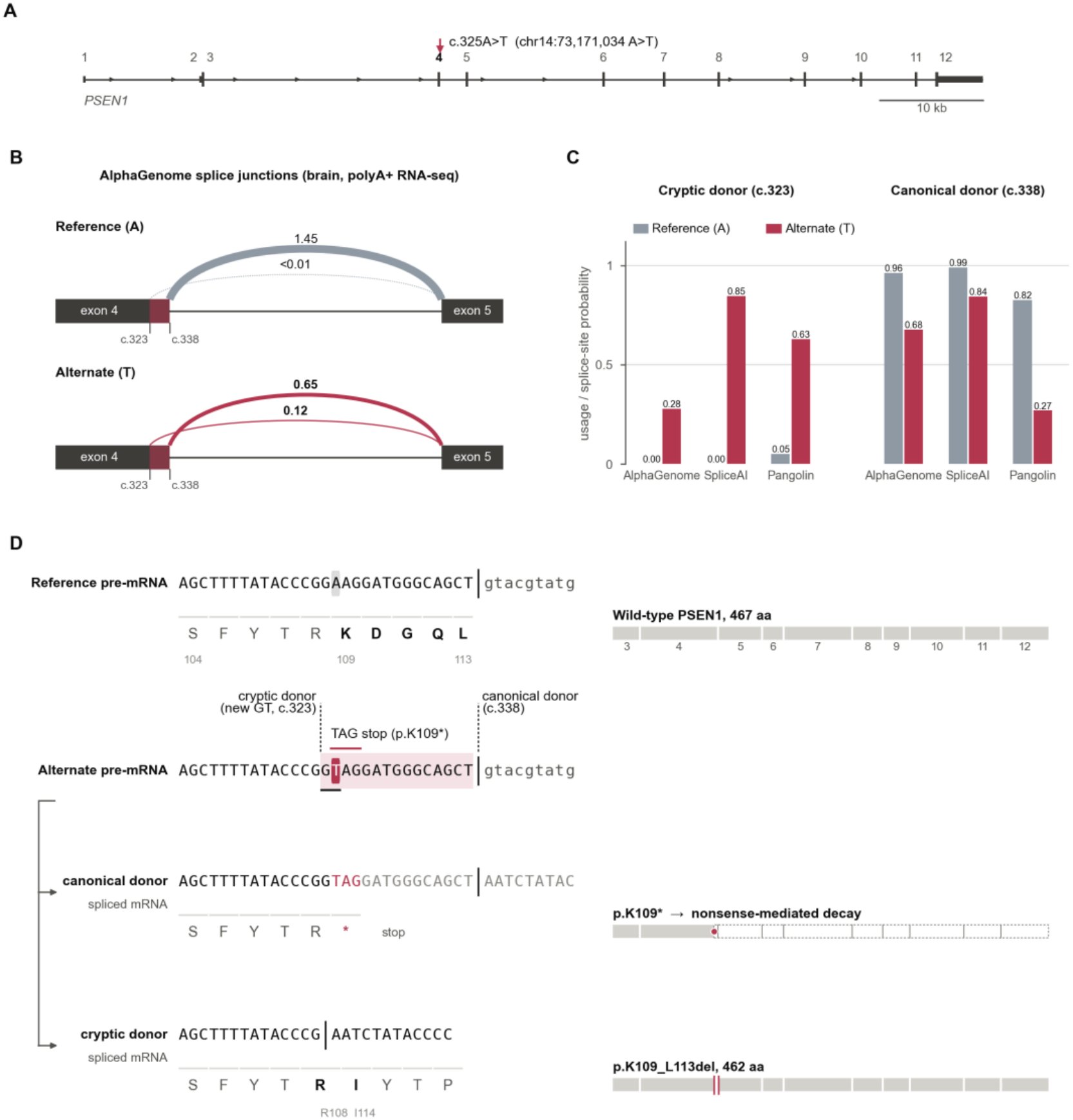
*PSEN1* c.325A>T creates a cryptic splice site. A) *PSEN1* gene model (GRCh38; ENST00000324501.10 / NM_000021.4) drawn to scale. The K109* variant (chr14:73,171,034:A:T, c.325A>T) is indicated with a red arrow. B) AlphaGenome-predicted splice-junction usage from exon 4 to exon 5 in brain polyA+ RNA-seq (ENCODE, UBERON:0000955). Line width is proportional to the predicted usage value. C) Predicted use of the cryptic (c.323) and canonical (c.338) exon 4 donors from three sequence-based splicing predictors. Y-axis units differ per model: AlphaGenome shows splice-site usage from the same run as B, SpliceAI shows donor probability, and Pangolin shows brain splice-site probability. All three tools predict a gain of the cryptic donor and a loss of the canonical donor. D) mRNA and protein outcomes of the c.325A>T mutation with canonical or cryptic splicing. The A>T mutation turns codon 109 into a TAG stop, leading to nonsense mediated decay with canonical splicing. Cryptic splicing with the new c.323 GT splice site removes the stop, leading to an in-frame 5 amino acid deletion (p.K109_L113del).

Although the K109* mutation may lead to a premature stop codon and nonsense mediated decay in a fraction of the *PSEN1* mRNA produced by carriers of this mutation, a significant fraction of *PSEN1* mRNA with the c.325A>T mutation likely undergoes alternative splicing, removing 5 amino acids (p.K109_L113del) from the full-length protein, including the newly created stop codon (Figure 3D). This alternatively spliced mRNA would escape nonsense mediated decay and likely create a full-length protein product. Interestingly, the 5 amino acids removed by the cryptic splice site include sites previously linked to familial AD^22,23^, suggesting that this alternatively spliced *PSEN1* could act through similar mechanisms as known *PSEN1* missense mutations.

We reviewed our other high-confidence *PSEN1* LoF mutations to determine whether any others may escape their loss of function effect through alternative splicing. Only one other mutation (a 68 base pair frameshift insertion at 14:73148090) had a SpliceAI predicted score over 0.5, the default cutoff for predicted splicing effects (Supplementary Table 4). The predicted use of a cryptic donor inside the insertion, rather than the canonical exon 3 donor, removes the frameshift, leading to an in-frame but altered protein product (p.N24_Q29delinsK). As this variant was only carried by one individual, observed as healthy into his 40s, it is unclear whether it leads to early-onset dementia. Overall, while certain mutations in *PSEN1* that create altered PSEN1 protein may cause EOAD, true genetic LoF mutations, in which a frameshift or stop-gain in the mutated allele leads to nonsense mediated decay, appear not to cause EOAD.

The case of the *PSEN1* K109* variant presents a cautionary tale, illustrating that alternative splicing may be key to interpreting the effects of high-impact variants. Although the K109* variant was experimentally studied in two model systems, both designs inadvertently prevented the detection of alternative splicing events: the cell model used overexpression of *PSEN1* cDNA lacking introns, and the gene editing strategy used to create a mouse K109* mutation also included a synonymous c.324G>T mutation to aid in genotyping, which did not affect the K109* protein product but eliminated the GT motif at the cryptic splice site^16^.

Interestingly, a 1998 study of two autopsy-confirmed EOAD cases had previously claimed to identify a frameshift variant leading to premature truncation of the PSEN1 protein, providing what they argued was the first LoF mutation supporting a haploinsufficiency mechanism for *PSEN1* in EOAD^24^. However, further mechanistic characterization revealed that an alternatively spliced protein with a single amino acid insertion (p.L113_I114insT) was in fact responsible for the disease phenotype^22^. Combined, these two cases illustrate both that novel *PSEN1* FAD mutations identified as LoF are more likely to be in-frame splice isoforms, and that more generally, apparent nonsense or truncation mutations should be carefully screened for alternative splicing consequences^25,26^. Recent work suggests that approximately 1 in 175 annotated nonsense variants may escape NMD through manufactured splice rescue^26^.

*PSEN1*-mediated FAD seems to require production of an altered PSEN1 protein product. This is consistent with a toxic gain-of-function intrinsic to the mutant protein, such as alterations in APP processing that change the profile of Aβ peptides produced. While not examples of gene-level loss-of-function, these alterations could also be understood as a partial loss of normal function at the biochemical level, with impaired ability of γ-secretase to trim longer Aβ intermediates into shorter peptide species as one example^27^. An additional possibility is a dominant negative effect, in which the mutant protein interferes with the activity of wild-type PSEN1^1,28^; however, the existence of direct physical interactions between wild-type and mutant PSEN1 has been contested^29^.

Overall, our observation of numerous carriers of gene-level *PSEN1* LoF mutations over the age of 65 without documented dementia suggests that this class of variants is not a Mendelian cause of early-onset Alzheimer’s disease. It is unlikely that *PSEN1*-mediated FAD is caused by *PSEN1* haploinsufficiency; a single wild type *PSEN1* allele appears sufficient to carry out AD-relevant *PSEN1* functions. These findings provide a mechanistic rationale for pursuing allele-selective suppression of mutant *PSEN1* in FAD, using approaches such as antisense oligonucleotides or siRNA. The apparent tolerance of heterozygous *PSEN1* nonsense mutations in humans supports the potential of selectively targeting mutant *PSEN1*, while preserving wild-type allele expression, as a viable therapeutic strategy for *PSEN1*-mediated FAD.

## Methods

### Cohorts

#### Alzheimer’s Disease Sequencing Project (ADSP)

The Alzheimer’s Disease Sequencing Project is a National Institute on Aging-funded collaborative initiative that integrates genetic and phenotypic data from multiple well-characterized, ethnically diverse cohorts of individuals with Alzheimer’s disease and related dementias, as well as cognitively healthy controls, to investigate the genetic architecture of disease susceptibility and resilience. We analyzed WGS and WES data from the ADSP release 5, comprising 58,507 genomes from 67 contributing cohorts hosted at NIAGADS^30^. Sample relatedness was assessed using KING v2.3.55. MCI and AD diagnoses were made according to ADSP criteria, supported by clinical assessment and, where available, AD biomarkers and/or neuropathological confirmation.

#### UK Biobank (UKB)

The UK Biobank is a large, prospective, population-based cohort study of individuals aged 40– 69 years recruited across England, Scotland, and Wales between 2006 and 2010, with extensive longitudinal data on participants’ health, lifestyle, environmental exposures, genetics, and linked medical records. Whole-exome sequencing (WES) data were available for 454,787 UKB participants as described previously^31^. We defined last known age as the age at last visit to a UKB assessment center; many individuals had non-dementia ICD codes recorded after their last visit date, but we chose the more conservative definition of last assessment visit due to the possibility that a dementia diagnosis could be missed during an unrelated hospitalization. We searched for dementia diagnoses using field 42018 (Date of all cause dementia report).

#### BioVU

BioVU is Vanderbilt University Medical Center’s de-identified DNA biobank, comprising biological samples obtained from discarded blood collected during routine clinical care and linked to de-identified electronic health records to facilitate genotype–phenotype association studies. Through the Alliance for Genomic Discovery (AGD), a collaboration between Vanderbilt University Medical Center, Nashville Biosciences, Illumina, and industry partners, whole-genome sequencing was performed on 250,000 BioVU samples^32^. The resulting genomic data are linked to de-identified clinical records through Vanderbilt’s Synthetic Derivative. AD and dementia diagnoses were assigned using the relevant ICD-9 and ICD-10 codes. Last observed age was computed by subtracting the participant’s birth date from the date of their most recent ICD code.

#### All of Us Research Program (AoU)

The All of Us Research Program is a large-scale, longitudinal cohort study initiated by the National Institutes of Health to enroll over one million participants across the United States, with an emphasis on populations historically underrepresented in biomedical research, collecting comprehensive demographic, clinical, lifestyle, and genomic data to advance precision medicine. We used the v8 WGS release (N = 414,830). We searched for dementia diagnoses by querying EHR tables by concept ID 4182210 (Dementia). Last observed age was computed by subtracting the participant’s birth date from their most recent EHR code date, including visits, diagnoses, drug exposures, procedures, and deaths.

### Loss of function variant identification

Only variants predicted by LOFTEE^17^ to be high-confidence loss of function mutations in *PSEN1* were considered. Each variant was annotated with Ensembl VEP through the Ensembl REST API (v116, GRCh38), using the LOFTEE plugin and the HGVS, exon/intron numbering and canonical/MANE options. Only consequences on the canonical *PSEN1* transcript were kept (ENST00000324501.10; MANE Select, NM_000021.4; 467 amino acids, 12 exons). gnomAD v4.1 information was queried directly from the gnomAD GraphQL API by variant ID, because the VEP REST frequency fields missed some variants present in gnomAD. Three variants found in gnomAD v4.1 with a FILTER status other than PASS were removed. Additionally, two frameshift insertions located two base pairs apart (14:73170830:A:ACCTT and 14:73170832:A:ACC) were always identified in the same individuals and on the same sequencing reads, suggesting they represent a single insertion event that was split into two variant calls. Because the combined insertion is in-frame, we did not consider these variants to be LoF.

Variants whose consequences included “frameshift_variant” or “stop_gained” were considered high-confidence LoF (labelled “NMD” in Supplementary Table 2) when they passed LOFTEE’s 50-bp rule, meaning that the premature termination codon lies more than 50 nt upstream of the last exon-exon junction, so nonsense-mediated decay is expected. Those that failed the rule were labelled “Truncated”, as they are predicted to escape NMD and produce a truncated protein. Variants whose only consequences were “splice_donor_variant” or “splice_acceptor_variant” were considered splice variants.

### Splice prediction

Alternative splicing effects of the K109* variant (chr14:73,171,034:A:T) were predicted using three different models.

AlphaGenome predictions were obtained from the AlphaGenome API^32^ (Python client alphagenome 0.10.0), using the default ALL_FOLDS model (distilled from teacher models trained on all folds). Reference and alternate predictions were generated with predict_variant on a 1,048,576-bp (1 Mb) input interval centered on the variant. We requested the SPLICE_JUNCTIONS and SPLICE_SITE_USAGE outputs, restricted to brain (UBERON:0000955; ENCODE polyA+ RNA-seq).

SpliceAI (v 1.3.1) was run with TensorFlow 2.15.1 and Keras 2.15.0 on Python 3.9.6. Per-base acceptor and donor probabilities for each allele were obtained by running the five bundled SpliceAI models on the GRCh38 reference and alternate sequences (5,000 nt of context on each side with a ±50 bp scoring window), following the custom-sequence usage in the SpliceAI documentation, and averaging their outputs as in the command-line tool.

Pangolin^33^ (v 1.0.2) was installed from its GitHub repository and run locally with PyTorch 2.15.0 (Python 3.11). Pangolin’s standard scores report only the largest change across its four tissues (heart, liver, brain, testis), so we ran the brain P(splice) models directly. The three brain ensemble members (v2 weights) were applied to the GRCh38 reference and alternate sequences (±500 nt around the variant, with 5,000 nt of flanking context on each side, + strand), with outputs averaged across members as performed in Pangolin’s default scoring.

All other high-confidence LoF and splice variants were scored with SpliceAI v1.3.1 as described above, and the maximal delta scores (acceptor gain, acceptor loss, donor gain and donor loss), and the position of each affected site relative to the variant, were recorded (Supplementary Table 4). A variant was flagged as potentially splice-altering when its maximum delta score was above 0.5. Potential splice effects were modelled on the canonical *PSEN1* transcript: a gain moved the nearest annotated splice site of the same type to the new position, and loss at an annotated site caused that exon to be skipped, unless a gain in the same run replaced the site.

For high-confidence LoF variants, LoF was considered removed (“Splice_LoF_removed” = yes) when the predicted product rejoined the wild-type reading frame with no premature stop or frameshift. This occurred for exactly one variant (14:73148090:A:AGGTACAGTGTCTGTACTGTACGTACAGCCAGGTACAGTGTCTGTACTGTACTGTAGTGTACTGTACTG), a frameshift insertion 16 nt before the canonical exon 3 donor. As SpliceAI predicted a loss of the canonical donor and gain of a cryptic donor 1nt into the insertion, the resulting 15nt difference in the predicted spliced RNA would restore the reading frame; thus, alternatively spliced mRNA with this variant would be predicted not to undergo NMD.

## Data Availability

Data used in preparation of this manuscript can be obtained upon application at:
- UK Biobank (https://biobank.ndph.ox.ac.uk/showcase/)
- ADSP (https://dss.niagads.org/datasets/ng00067/)
- All of Us (https://workbench.researchallofus.org/)
Data access for the BioVU database is limited to Vanderbilt-affiliated investigators following an application process.

